# PD Union: An Automated Pharmacodynamic Modeling Framework Based on a Unified Mechanistic Skeleton and Machine Learning Assistance

**DOI:** 10.64898/2026.05.05.26352278

**Authors:** Songrui Du, Mengjie Fan, Dongyang Liu

**Author notes:** Correspondence: Dongyang Liu, Drug Clinical Trial Center, Peking University Third Hospital, Beijing 100191, China; Institute of Medical Innovation, Peking University Third Hospital, Beijing, China.

## Abstract

**Objective:** Conventional pharmacodynamic (PD) modeling workflows require manual model selection, repeated equation rewriting, and empirical parameter adjustment, resulting in limited automation, high cross-scenario migration costs, and insufficient reproducibility. This study aims to develop PD Union, a unified, automated, and interpretable framework for mechanistic PD modeling.

**Methods:** PD Union is built upon a unified continuous dynamical skeleton that organizes absorption and systemic exposure module, the receptor module, the drug input module, the first delay module, the primary pharmacodynamic function module, the primary pharmacodynamic state module, the downstream pharmacodynamic state module, the second delay module, the feedback module, the circadian modulation module, the biophase module, the direct effect module, the disease state module, the second PD axis first delay module, the second PD axis primary pharmacodynamic function module, the second PD axis primary pharmacodynamic state module, the second PD axis downstream pharmacodynamic state module, the second PD axis second delay module, and the second PD axis feedback module. A machine learning-based structure identification module is incorporated to recognize drug input modes and mechanism labels from population PK/PD time series, followed by constrained population parameter optimization, forming an integrated pipeline of structure identification, candidate generation, and parameter fitting.

**Results:** Validation was conducted at two levels. In standardized synthetic benchmarking across 14 representative single-endpoint scenarios, the structure identification model achieved an output mode accuracy(NRMSE) of 0.7600 and macro-average F1 of 0.6307; parameter fitting yielded an NRMSE mean of 0.146 and median of 0.117. In the unified reconstruction validation based on 15 population pharmacokinetics/pharmacodynamics (PK/PD) literature data, the mean NRMSE of PDUnion model for PD was 0.261, and the median was 0.228. Among the 15 studies, 14 performed better than the models provided in the original literature.

**Conclusions:** PD Union demonstrates that interpretable mechanistic modularization combined with machine learning-assisted structure identification is feasible for automated PD modeling. The framework provides an executable methodological foundation for unified, reproducible, and extensible mechanistic PD modeling, with potential applicability to multi-endpoint and complex disease-state modeling scenarios.

## 1. Introduction

### 1.1 Clinical and Translational Research Background

In clinical practice, even when the same dosing regimen is administered, substantial variability in therapeutic efficacy and adverse reactions is commonly observed across patients. How to achieve more precise drug administration based on patient characteristics, disease status, and temporal dynamics is a central question in precision medicine and translational research^1^. Addressing this question requires connecting drug administration, systemic exposure, biological response, and clinical outcomes into a computable and predictable quantitative relationship.

This is the core task of quantitative pharmacology, in which pharmacodynamics (PD) characterizes the quantitative relationship between drug exposure and biological response, serving as a critical link between pharmacokinetic processes, pharmacological mechanisms, and clinical decision-making^2^. Unlike pharmacokinetics, which primarily describes concentration changes, PD directly addresses biological responses that are simultaneously influenced by target binding, signal transduction delay, feedback regulation, circadian rhythms, disease progression, and individual variability. Therefore, PD is a very complex and difficult-to-standardize component in quantitative pharmacology.^3^.

To address the quantitative characterization of drug exposure-response relationships, traditional approaches primarily rely on mechanistic PD modeling frameworks, including classical model families such as direct effect models, indirect response models^4^, effect compartment models^5^, feedback models^6^, circadian models^6^, disease progression models^7,8^, and tumor growth inhibition models^9,10^. The strength of these approaches lies in strong physiological interpretability: modeling results can be traced back to well-defined mechanistic structures and readable parameters, which is why this approach has long been regarded as the gold standard in mechanistic PD research.

However, traditional approaches also have significant limitations. First, different PD scenarios often correspond to different model families, requiring researchers to repeatedly perform structural judgment, equation rewriting, and module reorganization, resulting in high migration costs across projects, endpoints, and disease scenarios. Second, model fitting is typically heavily dependent on empirical initial values, parameter boundary settings, and multiple rounds of manual trial-and-error; even when faced with similar scenarios, different modelers may reach different structural choices and fitting results. Third, as studies involving multiple endpoints, population data, and complex disease states continue to grow, the originally fragmented model families are increasingly difficult to unify and efficiently reuse within the same technical system.In the unified reconstruction validation based on 15 population pharmacokinetics/pharmacodynamics (PK/PD) literature data, the mean NRMSE of PDUnion model for PD was 0.261, and the median was 0.228. Among the 15 studies, 14 performed better than the models provided in the original literature.

### 1.2 Limitations of Existing Automation and Machine Learning Methods

In recent years, researchers have explored machine learning methods to improve PK/PD modeling efficiency, such as for feature extraction, covariate selection, parameter prediction, time-series classification, or structure search^11-14^. These methods provide important inspiration for automation, demonstrating that pattern information in complex biological time series can be identified through data-driven approaches^15,16^, thereby partially replacing the expert-dependent preliminary screening process in traditional workflows.

However, existing methods still suffer from two prominent problems. One category of methods leans toward pure data-driven approaches: although they can achieve high fitting accuracy in prediction tasks, they often fail to provide structural explanations consistent with pharmacological mechanisms, and their outputs are typically difficult to translate into clinically interpretable parameters or to be directly adopted by mechanistic researchers. ^13,17^The other category of methods retains mechanistic model shells but has limited automation, often only introducing auxiliary algorithms at isolated steps, without genuinely solving the problem of coherently connecting structure selection, candidate screening, and parameter fitting^18^.

Therefore, what the field urgently needs is not yet another black-box model that merely pursues numerical accuracy, but rather an automated PD modeling framework that can significantly lower the modeling barrier while maintaining mechanistic interpretability and clinical readability.

### 1.3 Research Objectives and Overall Approach

Against this background, this paper aims to establish an automated and interpretable methodological framework for mechanistic PD modeling, enabling structure recommendation and model construction—processes that have traditionally relied heavily on expert experience—to be performed within a standardized pipeline.

We specifically aim to achieve three objectives: first, to unify the expression of multiple classical PD mechanisms within a single continuous dynamical system, thereby reducing the inconvenience caused by repeated model selection and reconstruction; second, to leverage machine learning for intelligent recommendation of candidate structures, significantly lowering the modeling barrier and narrowing the structural search space; and third, to automatically fit optimal parameters to obtain the final PD model. Throughout this automated pipeline, the interpretive framework consistent with the current gold standard of mechanistic PD modeling is preserved, such that the final output structures and parameters retain pharmacological and clinical interpretive value.

To achieve These objectives, we construct the PD Union framework: built upon a unified mechanistic skeleton, it organizes modules including drug exposure, receptor binding, delay transduction, primary pharmacodynamic formation, feedback regulation, biophase, direct effect, disease state, the second PD axis first delay module, the second PD axis primary pharmacodynamic function module, the second PD axis primary pharmacodynamic state module, the second PD axis downstream pharmacodynamic state module, the second PD axis second delay module, and the second PD axis feedback module.Machine learning models are then applied to perform structure identification and candidate screening on standardized PK/PD time series, followed by parameter optimization, fitting diagnostics, and result selection under candidate structure constraints. This achieves mechanistic PD modeling through an intelligent pipeline with lower barriers, higher consistency, and greater generalizability, reducing the threshold for PD modeling, improving clinical efficiency, and fully supporting the demand for scalable predictive capabilities in precision medicine and translational pharmacology.

## 2. Method

### 2.1 Overall Framework of PD Union

PD Union is a unified, automated, and interpretable methodological framework for population pharmacodynamic modeling. Its core objective is to abstract recurrent mechanistic components in classical PD modeling into a uniformly organized set of composable mechanistic modules, enabling structure identification, candidate model assembly, parameter estimation, and model selection within a single dynamical parent system. In doing so, PD Union transforms the traditionally expert-driven, trial-and-error modeling process into a standardized, reproducible, and traceable pipeline.

The fundamental premise of PD Union is that different PD models should not be treated as mutually isolated model families, but rather understood as distinct mechanistic combinations along a shared pharmacodynamic axis. On this basis, multiple classical PD scenarios can be represented coherently within a unified framework, thereby enabling cross-scenario comparison, transfer, and automated recommendation. From an implementation perspective, PD Union comprises three interconnected layers. The first layer is the unified mechanistic skeleton, which defines the shared structural boundaries and modular organization of pharmacodynamic models. Built upon this skeleton, the structure identification module first identifies the most likely driving modes and mechanistic combinations from data, thereby narrowing the model search space. Subsequently, the parameter fitting module performs parameter estimation, model comparison, and diagnostic evaluation for candidate models within the unified skeleton. Within this framework, machine learning serves to assist structure search and priority ranking, while the final output remains a mechanistic model with clearly defined physiological and pharmacological interpretations.

The overall architecture of PD Union, encompassing all five components from problem formulation to real-literature validation, is illustrated in Figure 1.

**Figure 1.**
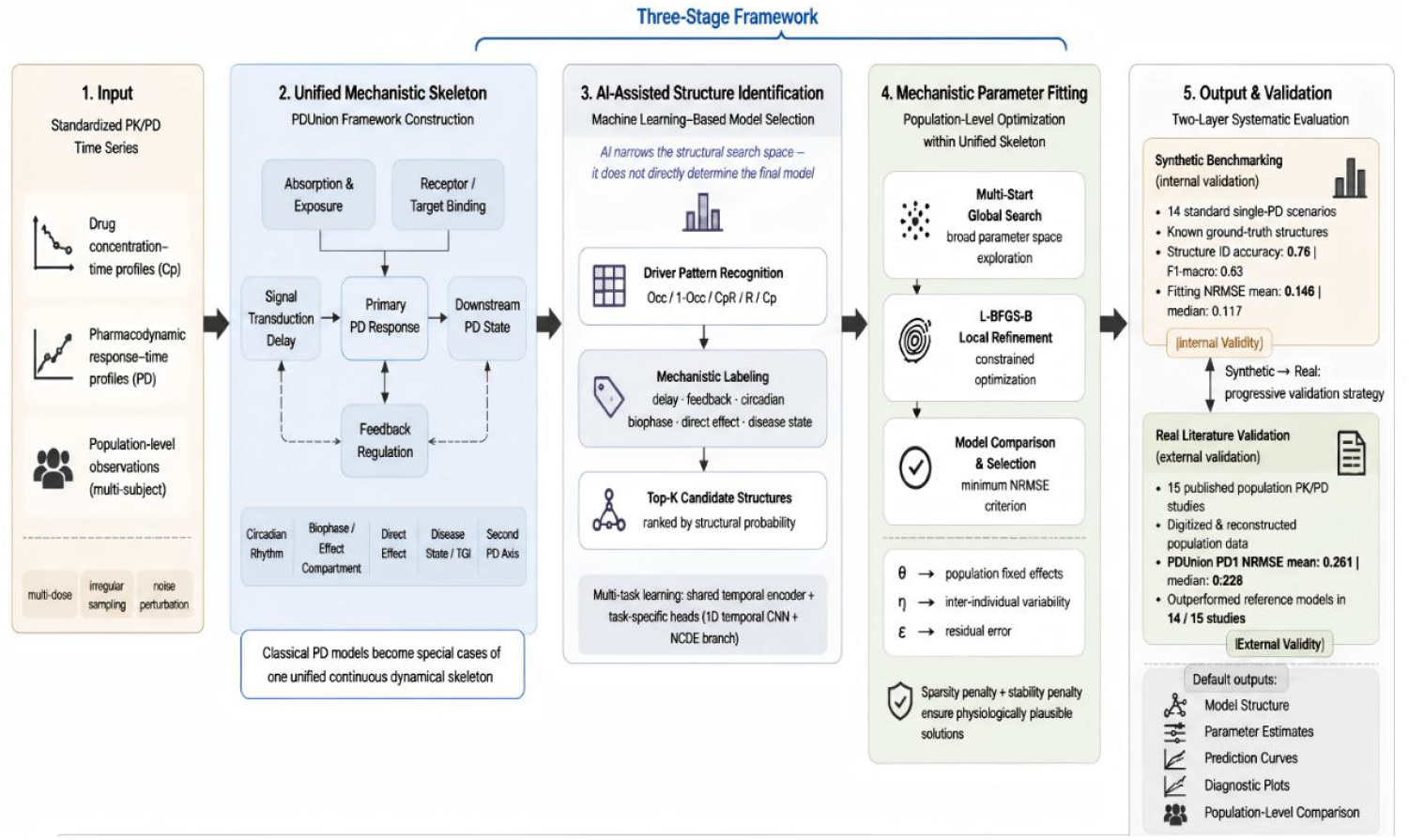
Overall framework of PD Union. Overview of the PDUnion automated pharmacodynamic modeling framework. The framework consists of five stages: (1)Input, standardized population PK/PD time series; (2) Unified Mechanistic Skeleton, integrating classical PD mechanistic modules into a single dynamical parent system; (3)AI-Assisted Structure Identification, multi-task machine learning for driving pattern recognition and candidate structure ranking; (4) Mechanistic Parameter Fitting, population parameter estimation via multi-start global search and L-BFGS-B refinement; (5)Output & Validation, evaluated through synthetic internal validation (NRMSE mean = 0.146) and external validation on 15 real publications (NRMSE mean = 0.261, outperforming reference models in 14/15 studies).

### 2.2 Design Principles of the Unified Mechanistic Skeleton

The unified mechanistic skeleton is the methodological core of PD Union. Based on the common biological logic of pharmacodynamic action, this skeleton predefines a hierarchically organized set of mechanistic axes, enabling different pharmacodynamic scenarios to be expressed under a consistent set of organizational principles. In other words, what PD Union unifies first is not specific parameter values, but the structured representation of the process by which drugs produce their effects.

The feasibility of this approach lies in the fact that most classical PD models, despite their apparent differences in surface formulation, are fundamentally organized around a limited set of relatively stable mechanistic questions: what drives the pharmacodynamic effect, whether the drug signal involves a delay, whether the effect arises directly or through state transitions, whether downstream propagation is present, and whether feedback regulation, circadian rhythms, or disease-state modulation should be considered. These questions define different mechanistic combinations rather than entirely independent model universes. Accordingly, organizing these recurrent mechanistic units into a unified skeleton does not weaken their original mechanistic interpretability; instead, it restructures traditional model formulations into a clearer, more coherent, and more verifiable structural system.

This design also offers two methodological advantages. First, the unified skeleton preserves the interpretive tradition of mechanistic PD modeling, ensuring that each invoked module corresponds to a well-defined pharmacological or physiological process, making it easier for researchers to understand the model origins and for reviewers to examine mechanistic assumptions and parameter meanings. Second, the unified skeleton provides a common foundation for subsequent automated modeling, allowing structure identification, parameter fitting, and model comparison to be completed under unified standards without requiring separate equation rewriting for each scenario, thereby significantly improving cross-scenario modeling consistency and reproducibility.

### 2.3 Modular Organization of PD Union

Within the same unified mechanistic skeleton, PD Union organizes the following modules according to the mechanistic steps that may be involved in the pharmacodynamic formation process: the absorption and systemic exposure module, the receptor module, the drug input module, the first delay module, the primary pharmacodynamic function module, the primary pharmacodynamic state module, the downstream pharmacodynamic state module, the second delay module, the feedback module, the circadian modulation module, the biophase module, the direct effect module, the disease state module, as well as the second PD axis first delay module, the second PD axis primary pharmacodynamic function module, the second PD axis primary pharmacodynamic state module, the second PD axis downstream pharmacodynamic state module, the second PD axis second delay module, and the second PD axis feedback module. Among these, the absorption and systemic exposure module characterizes the process of drug entry into the body and formation of systemic exposure; the receptor module and drug input module define the form in which the drug drives the subsequent pharmacodynamic system; the first delay module, second delay module, and feedback module characterize the time lags and regulation involved in drug signal propagation and pharmacodynamic recovery; the primary pharmacodynamic function module, primary pharmacodynamic state module, and downstream pharmacodynamic state module describe the formation of the pharmacodynamic effect and its transfer to downstream states; the circadian modulation module, biophase module, direct effect module, and disease state module characterize more complex real-world pharmacodynamic processes; and the second PD axis-related modules are used to express a second pharmacodynamic endpoint or a second response pathway in parallel within the same skeleton, with corresponding delay, pharmacodynamic formation, state propagation, and feedback regulation processes mirroring those of the first PD axis (see Figure 2).

**Figure 2:**
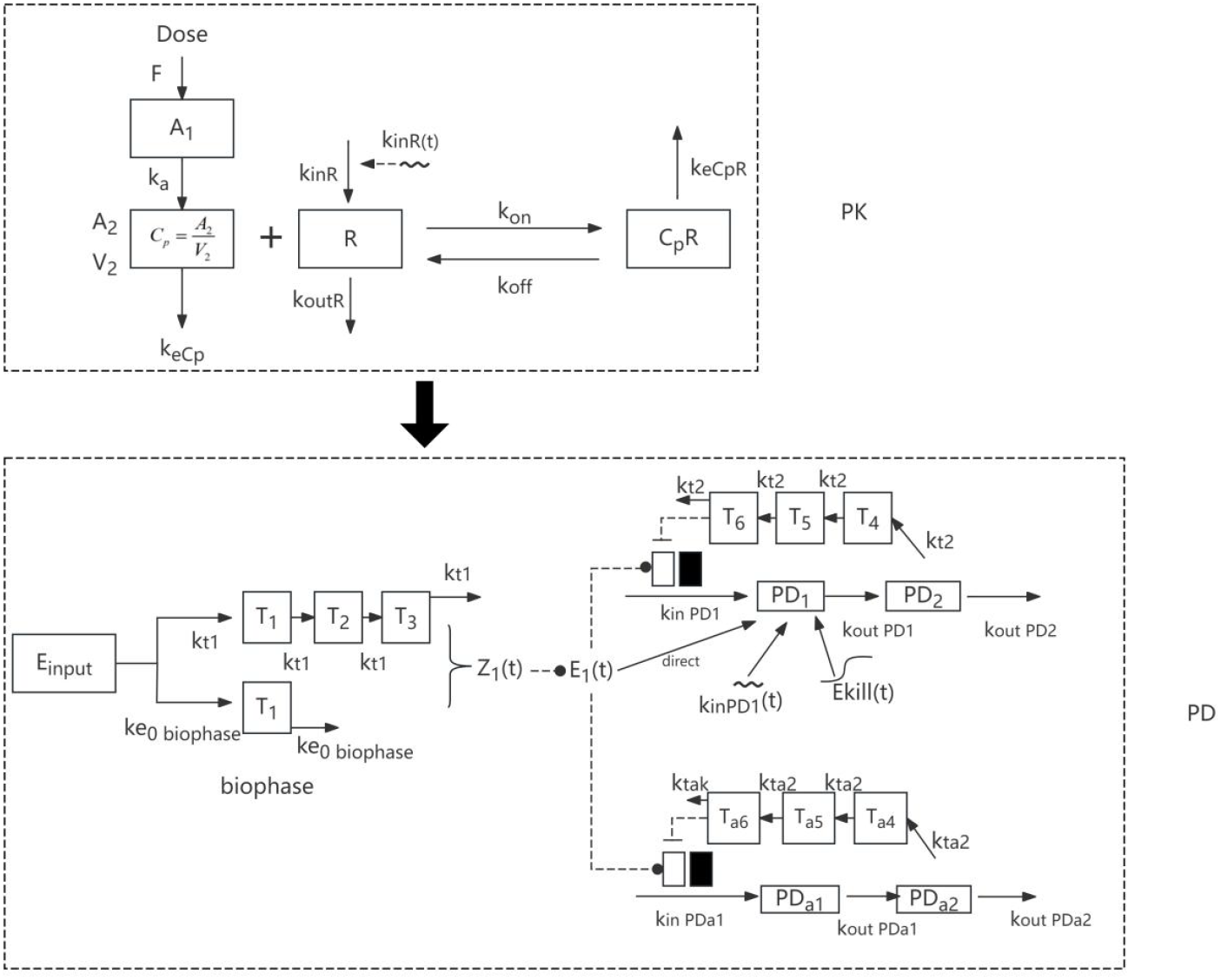
PD Union Framework.

During concrete modeling, these modules are selected, combined, and invoked within the unified skeleton according to the data characteristics and mechanistic assumptions of the current research problem. A specific PD model can be understood as a particular combination of these parallel modules for a given scenario. For a specific study, the key to model construction is no longer inventing a model from scratch but rather determining which modules within the unified skeleton should be activated, which should remain inactive, and how these modules should be combined.

### 2.4 Construction and Application of the Structure Identification Module

To improve the efficiency of structural judgment and accelerate the subsequent modeling process, we first constructed the structure identification module, which identifies the most probable pharmacodynamic driving modes and mechanistic combinations from data prior to formal parameter fitting. The role of this module is to provide the most likely candidate structures, thereby narrowing the search range for subsequent fitting and transforming the preliminary structural judgment, which traditionally relies mainly on researcher experience, into a data-driven recommendation process.

#### 2.4.1 Construction of Training Data

The training of the structure identification module is primarily based on simulated pseudo-data. The use of pseudo-data grounded in real cases is motivated by two main considerations. First, the number of PD cases directly available for model training is currently limited, particularly cases that simultaneously include raw time-series data, relatively complete parameter information, and relatively well-defined structural interpretations, making it difficult to support supervised learning for multi-module identification tasks. Second, the training objective of the structure identification module is not to replicate any specific study, but to enable the model to learn which mechanistic modules should be invoked; therefore, the key requirement for training data is to provide clear, reliable, and traceable ground-truth module labels.

This pseudo data is mainly used to train the structural recognition module to initially identify the time series features of different mechanism combinations, rather than directly replacing the real data to complete the final model selection. Since the pseudo data is generated under a unified mechanism framework, the driving method and mechanism module combination of each sample are known during generation. Therefore, it can provide clear and traceable supervisory labels. By systematically perturbing the parameter range, sampling scheme, and observation noise, the pseudo data can enhance the model’s ability to learn stable dynamic characteristics under different experimental conditions. However, the results obtained from the pseudo data training should be regarded as candidate structure screening or prior suggestions. The final model selection still needs to be comprehensively judged by combining the fitting performance of the real observation data, parameter identifiability, prediction verification, mechanism rationality, and external validation results.

#### 2.4.2 Multi-Task Training Strategy and Three Task Implementations

In terms of model implementation, the structure identification module adopts a multi-task learning framework consisting of a shared temporal representation layer and task-specific output heads^19^. It consists of a one-dimensional time encoder^20,21^.: extracting local dynamic morphological features from the Cp and PD time series. The output time features are then input into three task branches, which are respectively used to complete the recognition of drug input patterns, the identification of mechanism labels, and the provision of parameter prior suggestions.

The first task is drug input mode identification. The core of this task is to determine in what input form the drug drives the subsequent PD system. The five currently defined input modes are receptor occupancy-driven (Occ), unoccupied receptor-driven (1-Occ), drug-receptor complex-driven (CpR), free receptor-driven (R), and drug concentration-driven (Cp). These five modes correspond to different mechanistic assumptions about how the drug action signal enters the PD module, and therefore directly influence the construction direction of subsequent candidate structures. This task is modeled as a supervised multi-class classification problem. The model first uses the shared temporal representation layer to extract dynamic features from Cp and PD time series, with the one-dimensional temporal encoder learning local morphological information; the extracted features are then fed into a five-class classification output head to assign normalized probabilities to each of the five input modes, thereby completing drug input mode identification.

The second task is mechanism label identification. The core of this task is to determine which mechanistic modules within the unified skeleton should be activated and which should remain inactive. The currently identified mechanism labels include delay, feedback, circadian rhythm, direct action, biological phase, and disease state. These are essentially all module-switching determination problems. The remaining modules within the unified framework are always active components and are defaulted to be enabled in all candidate structures. In terms of algorithmic implementation, this task is modeled as a supervised multi-label classification problem sharing the same temporal representation layer as the first task. The model first extracts dynamic features from Cp and PD time series via the one-dimensional temporal encoder, then feeds the shared features into a multi-label classification head to output the activation probability for each mechanism label independently. A threshold rule is applied to the predicted activation probability of each mechanism module: probabilities above 0.7 are set to active (1), those below 0.3 are set to inactive (0), and those between 0.3 and 0.7 are enumerated as both active and inactive. All combinations across uncertain modules form the candidate mechanism set, which is then paired with candidate driving modes and ranked to produce the final Top-K candidate structure set. Subsequently, based on the predicted mechanism label results, it determines which module combinations should be prioritized in the candidate structure generation stage and further generates a Top-K candidate structure set with ranking information. The role of the second task in the overall pipeline is to transform the mechanistic information contained in the time series into module switch judgments within the unified skeleton, directly serving the generation and ranking of subsequent candidate structures.

The third task is parameter prior suggestion. The core of this task is, after completing input mode identification and module switch determination, to further predict which reasonable range the key parameters associated with the current candidate structure should fall into, thereby providing more appropriate initial values and search boundaries for subsequent parameter fitting. In terms of algorithmic implementation, this task is modeled as a supervised continuous regression problem sharing the same temporal representation layer as the previous two tasks. The model first extracts dynamic features from Cp and PD time series via the one-dimensional temporal encoder, then feeds the shared features into a parameter regression head to predict the prior range or scale information of key parameters. During training, the traceable true parameter values from the pseudo-data generation process or their normalized representations serve as supervisory signals, enabling the model to learn the correspondence between different dynamic morphologies and parameter intervals. The role of this task in the overall pipeline is to narrow the parameter search range for subsequent candidate structure fitting, improve the rationality of parameter initialization, and reduce ineffective searches during numerical optimization.

In summary, driving mode identification constrains the basic form of drug action input, mechanism label identification constrains the module combinations that should be prioritized, and parameter prior suggestion constrains the parameter search region for subsequent fitting. After obtaining driving mode probabilities, mechanism label probabilities, and parameter prior suggestions, the system further combines these with the module logic constraints within the unified skeleton to generate a Top-K candidate structure set with ranking information, for further comparison and selection by the subsequent parameter fitting module.

### 2.5 Parameter Fitting

After generating the Top-K candidate structures with sorting information in the structural recognition module, PD Union subsequently performs parameter estimation for each candidate structure individually. Under a given structure, it identifies the parameter set that best explains the current data.

This study employs a parametric approach based on population pharmacokinetics. Once the candidate model structure is established, the population fixed-effect parameter ‘θ’ represents the typical parameter levels of the target study population under the current pharmacodynamic structure. Building upon this, ‘η’ denotes interindividual variability, describing how parameters of different subjects deviate from the population mean; while ‘ε’ represents residual error, capturing the random deviation between observed and predicted values that cannot be accounted for by structural or individual differences. Consequently, the final fitted parameters constitute a parametric framework for population modeling, defined by ‘θ’ as the population mean, ‘η’ for interindividual variability, and ‘ε’ for residual error.

Parameter estimation is performed independently within each candidate structure and follows a unified numerical optimization process. The current approach employs a two-stage strategy: “multi-start global search + L-BFGS-B local refinement.” First, multi-start initialization is conducted within a well-defined parameter space to enhance global accessibility under complex objective functions; subsequently, L-BFGS-B is applied to locally refine candidate feasible solutions, yielding stable and interpretable parameter estimates. This strategy essentially continues the conventional approach in traditional mechanism modeling—first identifying a feasible parameter region and then performing local high-precision optimization—only transforming the previously manual trial-and-error process into automated execution within a unified program.The objective function corresponding to parameter estimation consists of three components:

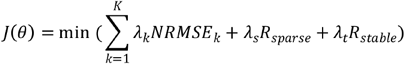

The first part presents the normalized root mean square error for each efficacy endpoint, which measures the deviation between model predictions and observed data under the current parameters.

The second part consists of a sparsity penalty term, which imposes L1 regularization constraints on the gating parameters related to the intensity of mechanism effects in the candidate structure, thereby reducing the influence of mechanisms lacking sufficient data support to near zero during the fitting process. The third part is a stability penalty term, designed to mitigate unstable dynamic behaviors such as non-physiological negative parameters, abnormal reentrant activities, and excessive oscillations.

During the candidate structure comparison phase, the system calculates the normalized root mean square error (NRMSE) for each candidate structure under its optimal parameters, with the minimum NRMSE value serving as the criterion for identifying the optimal structure. Ultimately, the candidate structure with the smallest NRMSE and its corresponding optimal parameters jointly constitute the pharmacokinetic model for the target pharmacodynamic study.

### 2.6 Final Model Determination and Diagnostic Output

After the final model is determined, the system further outputs the time-series fitting curves, observed-versus-predicted scatter plots, and residual diagnostic plots corresponding to that model, demonstrating the model’s reconstruction of the pharmacodynamic dynamic process and assisting researchers in checking for obvious systematic biases or abnormal dynamic behavior. The role of graphical diagnostics in this pipeline is primarily result presentation and auxiliary verification, while also allowing human experts to review the final results.

### 2.7 Result Evaluation

Result evaluation is conducted at two levels. The first level uses standardized population pseudo-data. This data is derived from Monte Carlo simulation samples under the unified mechanistic skeleton, divided into training, validation, and test sets for training and evaluating the structure identification model, and also for evaluating the recovery capability of the parameter fitting model under known ground-truth conditions. Since the driving modes, module switch combinations, and parameter true values are known at the time of generation, the focus of this evaluation level is not only to assess whether the structure identification model can correctly identify the five drug input modes and corresponding mechanism modules, but also to verify whether the combined structure identification and parameter fitting pipeline can recover the correct structure and retrieve reasonable parameters under known true-value conditions.

The second level uses real population PK/PD literature uniformly reconstructed data. Specifically, PD Union’s practical applicability is further evaluated on data reconstructed from 15 real population PK/PD publications. Unlike the first evaluation level, which mainly verifies the method’s identification and recovery performance under known true-value conditions, the focus of this level is to assess the framework’s transferability to real research data, that is, whether structure identification results can effectively support subsequent parameter fitting to ultimately yield pharmacodynamic models with good fitting performance and interpretable population parameters. The two levels of evaluation correspond respectively to verification of methodological correctness under known true values and verification of methodological usability in real application scenarios, collectively providing a systematic evaluation of PD Union’s performance.

## 3. Result

### 3.1 PD Union Model Construction Results

This study first completed the construction of the PD Union unified mechanistic skeleton. The unified mechanistic pharmacodynamic model skeleton includes the absorption and systemic exposure module, the receptor module, the drug input module, the first delay module, the primary pharmacodynamic function module, the primary pharmacodynamic state module, the downstream pharmacodynamic state module, the second delay module, the feedback module, the circadian modulation module, the biophase module, the direct effect module, the disease state module, the second PD axis first delay module, the second PD axis primary pharmacodynamic function module, the second PD axis primary pharmacodynamic state module, the second PD axis downstream pharmacodynamic state module, the second PD axis second delay module, and the second PD axis feedback module. The specific equations are presented as follows.

The absorption and systemic exposure module characterizes the process by which the drug enters the body and forms systemic exposure. The following equations are used to describe absorption and systemic exposure:

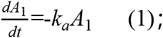

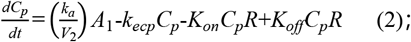

The receptor module characterizes the binding and dissociation of the drug with its target, the resulting changes in drug-receptor complex, and the turnover process of the receptor itself^22^. The following system of equations is used to describe these processes:

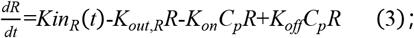

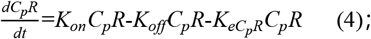

This set of equations describes the generation and elimination of receptors, as well as the processes of drug-receptor binding, dissociation, and complex formation/interaction.

The drug input module is used to characterize how the upstream dosage drives the downstream pharmacological effect.

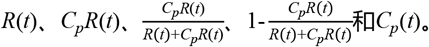

The first delay module describes the lag and smoothing process involved in the transmission of the drug action signal to the pharmacodynamic layer^23^:

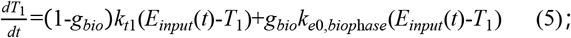

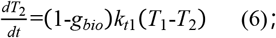

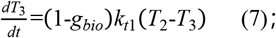

The primary pharmacodynamic function module maps the drug concentration in the effect compartment to the effective action intensity of the primary pharmacodynamic state. The specific functional representation is as follows:

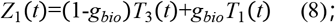

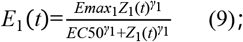

The primary pharmacodynamic state module describes the generation and elimination of the primary pharmacodynamic state along the first PD axis^4^.

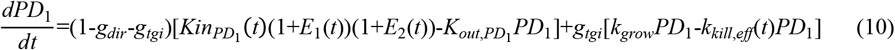

The downstream pharmacodynamic state module describes the transmission of the primary pharmacodynamic state to the downstream pharmacodynamic state:

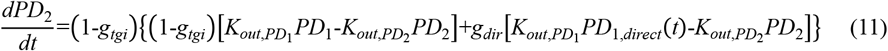

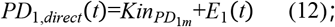

The second delay module describes the delayed transmission process of downstream pharmacodynamic changes prior to the formation of feedback:

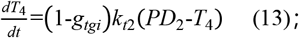

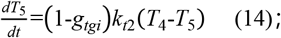

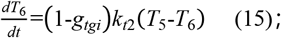

The feedback module performs the calculation of the feedback quantity (feedback intensity) through the first PD axis feedback function E2(t):

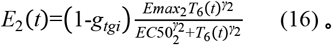

The rhythm modulation module describes the periodic fluctuations at the receptor generation end and the primary pharmacodynamic state generation end:

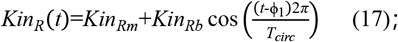

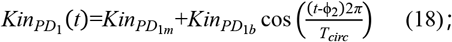

The disease state module describes the natural progression of the disease state quantity itself and the regulatory process of drugs on the disease state^7,8^, or the natural growth of tumor burden and the process of drug inhibition or killing of tumor burden^24^. The following auxiliary quantities are used for characterization:

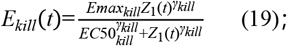

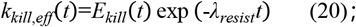

The second PD axis related modules:

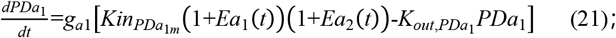

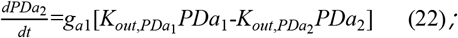

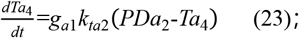

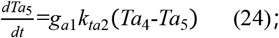

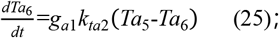

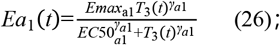

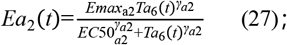

The meanings of the aforementioned specific parameters are detailed in Table S1. Based on equations (1)–(27), the unified mechanistic framework has integrated the core pharmacodynamic mechanisms throughout the entire process within a single continuous kinetic matrix.

### 3.2 Structure Identification Model Results

After completing the construction of the unified mechanistic skeleton, this study further established the structure identification model for identifying drug input modes and module switch combinations from population time series. Results show that PD Union is already capable of stably recovering key structural cues from time series. Based on the observable mechanism label version results, the model achieved an E_input_mode identification accuracy of 0.8769 on the test set, with multi-label identification performance of F1-micro 0.8481 and F1-macro 0.8300, demonstrating that the model can not only distinguish major driving forms but also relatively stably identify mechanism labels such as delay, feedback, and circadian rhythm.

Upon further extension to five drug input modes, the model now supports five driving forms including Occ, 1-Occ, CpR, R, and Cp. The current five-mode version achieved a test set identification accuracy of 0.7907 and F1-macro of 0.6368. This indicates that the output of the structure identification model is no longer limited to a single label judgment but can further provide driving mode probabilities, mechanism label probabilities, and a Top-K candidate structure set with ranking information, providing direct input for subsequent parameter fitting.

### 3.3 Parameter Fitting Results

After the structure identification model provided candidate structures, PD Union further completed population parameter estimation within the unified skeleton. In the current standardized population pseudo-data validation, we constructed a population study library covering 22 representative mechanistic prototypes. This library comprises 40 population studies in total, each of which may contain multiple study instances with different dosing regimens or sampling designs; across all 40 studies, there are 76 study instances in total, encompassing 4560 simulated subjects in aggregate. Batch fitting was completed across all studies, and all runs completed successfully, demonstrating that the framework already possesses batch population modeling capability To ensure comparability across studies, the current batch statistics uniformly use PD1 NRMSE as the primary indicator. Overall, the 40 studies yielded a PD1 NRMSE mean of 0.435647, median of 0.264221, minimum of 0.130471, and maximum of 0.939289. Further stratification shows that basic-difficulty studies achieved a PD1 NRMSE mean and median of 0.204536 and 0.207337, respectively(NRMSE value less than 30 is considered acceptable.). These results indicate that PD Union can already relatively stably recover the main pharmacodynamic dynamics for basic population scenarios, while error increases substantially in high-difficulty studies with more complex mechanisms and weaker identifiability, although the pipeline itself remains operationally stable.

From representative fitting results, in multiple study types including hypertension-type, INR-type, feedback-type, biophase delay-type, and receptor occupancy-driven-type scenarios, the population fitting curves of the basic-difficulty group can follow the main dynamic trends reasonably well, with observed-predicted scatter points mostly distributed near the line of unity and residuals showing no obvious systematic bias. This indicates that the coupled process of the unified mechanism framework, structure identification, and overall parameter estimation can generate stable and interpretable overall fitting results in the current mechanism scenarios.

### 3.4 Real Population Literature Validation

The real population literature validation further demonstrates that PD Union’s performance is not limited to standardized pseudo-data. On uniformly reconstructed data from 15 real population PK/PD publications, PD Union achieved a PD1 NRMSE mean of 0.260993, median of 0.227779, minimum of 0.125734, and maximum of 0.401204; the corresponding reference models from the literature achieved a NRMSE mean and median of 0.277444 and 0.263215, respectively. Under unified comparison criteria, PD Union outperformed the reference model in 14 out of 15 publications, with an average improvement of 0.016451.

These 15 publications cover a wide range of population PK/PD problems including antibody-biomarker suppression, acid suppression, sedation scores, long-term glucose lowering, renal function impairment, thrombocytopenia, metabolic effects, and disease score improvement. This demonstrates that PD Union is not effective only for a single type of templated study, but can transfer to real population modeling tasks involving different drugs, different endpoints, and different mechanistic backgrounds.

## 4. Discussion

This study reorganizes the fragmented PD model families using a unified mechanistic skeleton. For a long time, PD modeling in quantitative pharmacology has accumulated a large number of classical models, but these models have typically existed in a scattered manner organized around different scenarios, endpoints, and research conventions, requiring researchers to repeatedly switch between different model families, rewrite equations, and reorganize parameter systems when facing new problems. The value of PD Union lies not in replacing all existing models, but in providing a unified representational parent system in which high-frequency mechanistic elements such as drug exposure, receptor action, delay transduction, feedback regulation and disease state can be organized and invoked within the same system. For precision medicine and translational research, the importance of this unification lies in its ability to transform originally scattered empirical modeling activities into more standardized and reproducible quantitative pharmacology workflows, thereby enhancing the capacity of quantitative pharmacology to support predictive research.

This study achieves a combination of machine learning-based automated modeling and interpretability. Many existing automated methods emphasize predictive accuracy or time-series fitting capability but often fail to preserve mechanistic structure and parameter meanings; while traditional manual modeling, though highly interpretable, is heavily dependent on expert experience and iterative trial-and-error, limiting efficiency and reproducibility. The approach taken by PD Union is not to directly replace mechanistic modeling with black-box models, but rather to have machine learning handle structure identification and candidate ranking^25^, while the unified mechanistic skeleton handles parameter estimation and mechanistic expression. Consequently, the models ultimately produced are still consistent with the current gold standard PD models, with an interpretive logic aligned with the current gold standard pipeline for population PK/PD modeling. In other words, the automation in PD Union is not a departure from the existing methodological system but rather, while preserving the interpretive tradition of quantitative pharmacology, significantly lowers the barriers to structure search and parameter initialization.

This study also represents an organizational paradigm of interpretable mechanistic modularization combined with machine learning-assisted structure identification and mechanistic parameter refinement. From this perspective, the potential value of PD Union extends beyond pharmacodynamics itself. Any dynamic modeling task that simultaneously requires physiological or clinical interpretation and involves multi-mechanism combinations and structure selection problems could potentially benefit from this approach, such as more general physiologically-based mechanistic models, disease progression models, or automated construction of multi-scale biological models^18^. For this reason, the contribution of PD Union lies not only in delivering a currently operable automated PD modeling framework, but also in providing a transferable methodological prototype for the automated organization of biological and pharmacological models.

At the current stage, we have established a basic framework consisting of a unified mechanistic skeleton, structure identification, population parameter estimation, and real-literature validation, demonstrating that the path of interpretable mechanistic modularization combined with machine learning-assisted structure identification and mechanistic parameter refinement is feasible in pharmacodynamic modeling. Future work will proceed in two directions. The first is to gradually extend the current framework, which primarily covers PD scenarios, to more general biological and physiological model construction tasks, enabling it to handle more complex problems in disease progression, signal transduction pathways, multi-scale regulation, and systems biology. The second is to further integrate the framework with agent systems, creating a more complete automated closed loop that encompasses model construction, candidate structure comparison, parameter initialization, diagnostic plot generation, result verification, and report output^26^. If these directions can be continuously pursued, the significance of PD Union will no longer be limited to a single modeling tool, but could potentially develop into an intelligent working platform for interpretable biological modeling.

## 5. Conclusion

This paper presents PD Union, an automated pharmacodynamic modeling framework based on a unified mechanistic skeleton and machine learning assistance. The framework implements an integrated automated modeling pipeline of mode identification, candidate structure generation, and parameter fitting within a unified continuous dynamical system. Results from standardized synthetic scenarios and validation on 15 real-literature population datasets demonstrate that PD Union can maintain the interpretability of mechanistic models while significantly improving the consistency and automation of cross-scenario modeling. The framework not only provides an executable methodological foundation for unified, automated, and extensible mechanistic PD modeling, but also offers a methodological reference for the automated organization of other interpretable physiological dynamical models.

## Data Availability

All data produced in the present study are available upon reasonable request to the authors.

## Notes

### Competing Interest Statement

The authors have declared no competing interest.

### Funding Statement

This study did not receive any funding

### Author Declarations

The article utilized publicly available data from 15 real documents or reconstructed data based on them. The corresponding articles are all publicly accessible. Below are their sources： (1)DOI: 10.1002/cpt.70173 (2)DOI: 10.1093/jpp/rgaf123 (3)DOI: 10.1097/FTD.0000000000001310 (4)DOI: 10.1111/cts.70404 (5)DOI: 10.1111/pan.70050 (6)DOI: 10.2147/DDDT.S533428 (7)DOI: 10.1111/jdi.70034 (8)DOI: 10.1002/bcp.70477 (9)DOI: 10.1002/psp4.70181 (10)DOI: 10.1111/cts.70388 (11)DOI: 10.1002/psp4.70209 (12)DOI: 10.1002/psp4.70210 (13)DOI: 10.1007/s11095-026-04028-0 (14)DOI: 10.1007/s40262-025-01595-0 (15)DOI: 10.1007/s40262-025-01598-x

